# Validation and Prospective Testing of a High Sensitivity, Quantitative Analytic Assay for HER2 on Histopathology Slides

**DOI:** 10.1101/2025.05.06.25327097

**Authors:** Nay N.N Chan, Patricia Gaule, Julia Benanto, Liam Scott, Charles J. Robbins, Mengni He, Katherine Bates, Revekka Khaimova, Daniel C. Liebler, Regan Fulton, David L. Rimm

## Abstract

**Purpose:** The recent approval of antibody drug conjugates (ADCs) targeting HER2 (like Trastuzumab Deruxtecan or T-DXd) have led to challenges for the immunohistochemical (IHC) companion diagnostic test since the test was optimized for gene amplified levels of HER2. Here we develop and validate an objective test for low level HER2 expression toward more accurate selection of patients for T-DXd.

**Experimental Design:** We validated the high-sensitivity HER2 assay using a mix of the requirements for an IHC assay and that of a ligand binding assay. Then we prospectively tested it on 316 core biopsy specimens received by Yale Pathology Labs from August 2022 to August 2023

**Results:** Using a 40-case breast cancer tissue validation set, we find very high accuracy and precision with a coefficient of variation below 10% and define a reportable range for the assay in amol/mm^2^. These prospective cases show the dynamic range of HER2 expression, but also the discordance of pathologist scores with quantitative measurements, especially in the low range of HER2. We find that 6% of the cohort was below the limit of detection of this more sensitive assay while 71% of the IHC=0 cases were above the limit of quantification. Efforts are underway to determine a possible threshold expression level required for T-DXd response.

**Conclusion:** This assay validation study provides a method for accurate, objective measurement of HER2 and has the potential to improve selection of patients for T-DXd or similarly targeted ADC therapies in the future.

**Translational Relevance:** Antibody Drug Conjugates (ADCs) represent a novel class of targeted therapy, but in some ADC approvals the target is not assessed due to the lack of an objective and reproducible method for measurement of protein on slides. This paper describes, validates and tests a new fully quantitative method for ADC target assessment on histology slides.

## INTRODUCTION

Over the last few years targeted therapies (antibodies) conjugated to toxic payloads (called antibody drug conjugates or ADCs) have been proven to be effective therapy for advanced breast cancer and other tumor types. Although ADCs have been around for many years^1^, recent work with HER2 targeted ADCs has been particularly successful. This may be due the large dynamic range of expression of this protein or perhaps the incorporation of a successful unconjugated antibody therapeutic, trastuzumab. This therapeutic is effective for between 10% and 20% of breast cancers that have amplification of the *ERBB2* gene, where patient eligibility to receive trastuzumab is determined by a companion diagnostic assay that assesses the level of HER2 or *ERBB2* gene amplification in breast cancer cells.

Initially, a new HER2 specific ADC, trastuzumab deruxtecan (T-DXd or Enhertu), was shown to be highly effective in advance breast cancer cases with high levels of HER2 (Destiny Breast 01)^2,3^. This success triggered further trials of this ADC in patients with low-HER2 (Destiny Breast 04)^4^, and most recently in patients with ultra-low HER2 (Destiny Breast 06)^5^. The determination of HER2 levels was done using the same immunohistochemistry (IHC) assay that is used for trastuzumab. In the newer trials the efficacy was lower than in the early trial with high HER2 expressing patients, suggesting that more target was associated with better response. This observation of association with target and response could only be made by comparing trials except for the DAISY trial^6^which had cohorts with different levels of HER2 but was under-powered for target level comparison. However, the publication of the Destiny 6 trial^5^assessed target levels (IHC=1 vs IHC=2) as a function of response and showed that the IHC=2 cases were almost twice as likely to benefit, compared to IHC=1 (Hazard Ratio for IHC 2 = 0.43 and for IHC 1 = 0.74). This finding emphasizes the importance of assessment of target levels in patient tumors.

The IHC assays for assessment of HER2 protein were designed and tuned to separate *ERBB2* gene amplified specimens from unamplified specimens and thus is most effective in tumors with a lot of protein (millions of molecules per cell). While this has been a highly effective companion diagnostic test, its repurposing for low- or ultra-low HER2 is unfortunate. Chromogenic IHC has a limited dynamic range and thus using the existing assay for low-HER2 is like trying to weigh a mouse on a scale built for elephants^7^. The result of this approach has caused the pathologist to be asked to do the task beyond the capability of the human eye, to distinguish IHC=1 (patient gets T-DXd) from IHC=0 (patient does not get T-DXd). Studies have shown that pathologists will be between 26% and 85% concordant in assessment of this difference^8-11^, which means that whether or not a patient gets drug could depend on their pathologist rather than the biology of the tumor.

To address this issue, we have built an analytic assay that measures HER2 in units of amol/mm^2^ that removes most of the subjective aspects of HER2 assessment^12^. This assay, called high sensitivity HER2 (HS-HER2) has optimized conditions such that most cases are above the limit of detection (94%), yet the assay is in the correct dynamic range to distinguish undetectable HER2 from low levels of HER2 (like that seen in normal breast ducts).

Validation of IHC testing uses a standard protocol, initially published by Fitzgibbons et al in 2014^13^for analytic validation of IHC based on a systematic literature search resulting in Guidelines Statements as opposed to analytic formula as seen in laboratory assays such as the ligand binding assay (LBA). This is because the practice of IHC assessment is the subjective reading by a pathologist, not calibrated objective measurements as seen in an LBA. Based on the literature, the IHC guidelines included two recommendations, one for non-predictive assays and one for predictive assays. The recommendations included 20 specimens, 10 positive and 10 negative for non-predictive assays showing this number was sufficient for 90% concordance. For predictive assays the authors recommended a 40-case validation with 20 positive and 20 negative cases. This guideline was adopted by the College of American Pathologists (CAP) and is currently the standard used in evaluation of IHC labs for CAP inspections of CLIA labs.

While this standard served the medical/pathology community well for subjective, pathologist-read assays, as technology has advanced allowing objective quantitative assays for proteins on histopathology slides, this approach is no longer fit-for-purpose. The assays we have described and use in this work are, in some ways, more like LBAs (Ligand Binding Assays), than IHC. Thus, for this first pass as validation of a truly analytic assay to measure protein on histopathology slides, we have combined criteria for IHC with criteria for LBAs to produce a rigorous approach to validation of this new quantitative assay. Here we show construction of a calibration cell line microarray (CMA) and its usage as a standard for a 40-case analytic validation (supplemental figure 1). We then use this assay for a prospective assessment of 316 cases to determine the dynamic range of expression of HER2 protein (in attomoles/mm^2^) on histopathology slides in a real-world, academic lab setting.

## MATERIALS AND METHODS

### Study population and sample collection

Two sets of whole tissue section slides were accrued for this study. The first was the validation set composed of 40 breast cancer cases with a range of HER2 expression (IHC 0, 1+, 2+/FISH-, 2+/FISH+ & 3+) collected from Yale Pathology Tissue Archive between 2018 and 2020. The second cohort was a prospective case collection, accrued by collecting serial sections of core biopsies from breast invasive cancer patients received by Yale Pathology Labs at Yale New Haven Hospital between 2022 and 2023. The study was approved by the Yale Human Investigation Committee protocol #9505008219 and conducted in accordance with the Declaration of Helsinki adopted by the world medical association. The accrual included 322 cases, of which 6 cases were excluded due to insufficient tumor area, resulting in 316 patients suitable for analysis. Clinical information was obtained from the pathologist final reports in CoPath: a comprehensive anatomic pathology information system at Yale New Haven Hospital. Retrieved information included estrogen receptor (ER), progesterone receptor (PR) and HER2 status based on ASCO/CAP guidelines for scoring^14,15^.

### Cell line Microarray (CMA) construction

A calibrator cell line microarray (CMA; internally called CMA 625) was constructed using 9 cell lines with different HER2 and TROP2 expression levels, including JURKAT #HTB-152, TT #CRL-1803, DLD-1 #CCL-221, VCaP #CRL2876, BT20 #HTB-19, T47D #HTB-133, ZR-75-1 #CRL-1500, BT483 #HTB-121, and A431 # CRL-1555. The cell lines were procured directly from ATCC (American Type Culture Collection) and used without further authentication. Individual cell lines were cultured in accordance with the standard methods provided by ATCC. Cell pellets were produced by culturing cells to the peak of log phase (threshold of stationary phase), then frozen and shipped to Protypia (now Inotiv) for LC-MS/MS for HER2 and TROP2 protein quantity (data provided in attomoles/ug total protein). A second cell pellet from the same prep of each cell line was stored in 70% EtOH and shipped to Array Science LLC for CMA production. Four master blocks were constructed with an expected yield of 250-300 consecutive histology sections per block. Twofold redundancy of 9 cell lines were arranged in four rows randomly as shown in supplemental Figure 2 and cell lines were prepared according to the QDAP standard operating procedure (SOP_QDAP_011_V1).

For the unit conversion, LC-MS/MS values (attomol/ug) of total protein concentration and cell count was provided by Inotiv, then the total protein amount per 1,000,000 cells (ug/10^6^ cells) was calculated. Cells of each core on CMA625 were counted and divided by total area of tissue core (mm^2^ /10^6^ cells) determined by QuPath software. To convert to ug/mm^2^, the total protein amount per 1,000,000 cells was multiplied by the total area of cells per 1,000,000 cells. Relative HER2 and TROP2 protein abundance as determined by LC-MS/MS was reported in attomol/ug of protein. To transform attomol/ug to attomol/mm^2^, the attomol/ug of both targets were multiplied by the ug/mm^2^ of each cell line. A detailed description of these methods previously used in the construction of first version of HS-HER2 standard CMA515 is available in Moutafi et.al 2022^12^.

### Validation of High Sensitivity-HER2 (HS-HER2) Assay

The FDA approved Roche/Ventana 4B5 assay is used as a comparator or the legacy assay. That assay is optimized to separate *ERBB2* gene amplified cases that express very high levels of HER2. While the assay is fit-for-purpose as designed, it is less sensitive than some other assays for low levels of HER2^8^ To build a high sensitivity assay for quantitative measurement of HER2, we first had to optimize the antibody concentration used in the assay for maximum signal to noise. We used the 29D8 antibody (CST) as described in the pilot study^12^ and used a similar technique for antibody optimization (although not described in detail in that work). To maximize the sensitivity of HER2 protein detection, a standardization tissue microarray (TMA) was built by using archival specimens including 80 FFPE breast carcinomas seen at Yale Pathology between 1998 and 2011, 10 breast cell lines controls (prepared at Yale), and 10 non-tumor breast tissue cores. Cases were arranged in columns according to their HER2 status by IHC and FISH. HER2 negative (IHC 2+ Not AMP, 1+ and 0) spots were evaluated. Determination of the optimal concentration to maximize antibody sensitivity was performed as previously described^16^The titration curve uses the top 10% of signal (true signal) and the bottom 10% of signal (non-specific binding or noise) to determine the signal to noise (S/N) ratio of the antibody over a two log fold change in primary antibody concentrations. All other factors remained constant to remove bias. Using the HER2 standardization TMA, 7 concentrations of primary antibody were tested over 2 experimental runs. The results of those 7 concentrations are plotted and shown in supplemental figure 3. A signal to noise peak was observed at 1 ug/ml thus determined to be the optimal concentration of the HER2 primary antibody.

### Quantitative immunofluorescence (QIF) staining by HS-HER2 Assay

Due to the high variability^17^and insufficient sensitivity of conventional IHC tests^7^ using in the stratification of HER2-low patients, we recently developed a new singleplex quantitative immunofluorescence assay called High Sensitivity HER2 (HS-HER2) assay and validated as a CLIA/CAP certified laboratory developed test (LDT) in our clinical Quantitative Diagnostics in Anatomic Pathology (QDAP) CLIA lab at Yale. This HS-HER2 assay allows accurate determination of HER2 protein levels in tumor samples in attomoles/mm^2^. To quantify HER2 protein in 316 breast cancer core biopsies, the HS-HER2 assay was performed on a fully automated research stainer platform (Leica BOND RX autostainer, Leica Biosystems, CA, USA) with designated Leica BOND reagents.

In brief, CMA and tissue slides were offline baked at 60°C for at least 1 hour prior to autostaining. By loading one control CMA slide together with every 9 tissue slides in each BOND slide tray, HS-HER2 QIF staining was carried out by the following protocol; deparaffinization with BOND dewax solution (AR9222), antigen retrieval with BOND HIER epitope retrieval solution 2 (AR9640) at 97°C for 20 minutes, blocking with ReadyProbes Endogenous HRP & AP blocking solution (R37629, Invitrogen) for 10 minutes and with BSA for 30 minutes, 1 hour-incubation with primary rabbit monoclonal HER2 antibody (clone 29D8, 2165, IgG, Cell Signaling) at optimal concentration of 1ug/ml mixed together with 1:100 concentration of pan-CK (Clones AE1/AE3,M3515, Dako), amplification with Rabbit Envision+ System – HRP labelled polymer anti-Rabbit (K400311-2, Dako) mixed together with 1:100 dilution of a green-fluorescent Alexa Fluor 546 Goat-anti-Mouse IgG (H+L)Cross-Adsorbed Secondary Antibody (A11003, Invitrogen) for 1 hour, staining with 1:50 dilution of a red-fluorescent Tyramide Signal Amplification (TSA) Cyanin 5 (SAT705A001EA, Akoya Biosciences) for 10 minutes and nuclear staining with 1:500 dilution of a blue-fluorescent 4′,6-diamidino-2-phenylindole (DAPI) for 10 minutes. BOND TM wash solution (AR9590) was frequently used in wash steps between reagent incubation steps mentioned above. Subsequently, the excess DI water on each slide were wiped off carefully and coverslipped with ProLong Gold Antifade mounting reagent (P36930, Invitrogen).

### Digital Fluorescent Imaging and Image Processing

All QIF slides of control CMAs and breast cancer cases stained by the HS-HER2 assay were scanned at 20X magnification on the CyteFinder HT II multiplexed fluorescent imaging platform (RareCyte, Seattle WA). The CyteFinder imaging platform uses advanced, high speed, multi-channel systems with integrated machine learning algorithms, designed for digital pathology and clinical laboratories. We used the optimized fixed exposure times and specific filter sets for individual fluorophores i.e., DAPI for nuclear compartment, Alexa Fluor 546 for tumor CK masking and Cy5 for HER2 target.

Prior to viewing in QuPath for further analysis steps, the scanned images were subjected to the following preprocessing steps using python scripts: lossless compression of fluorescent images, conversion of QIF images to pseudo-DAB images (using method describe in https://github.com/serrob23/falsecolor), cleaning image file names, matching QIF images with pseudo-DAB images to create a sync-map file and creating batch-map file to match batch standards with QIF images. These additional files are used by Qymia for transferring annotations from pseudo-DAB images to QIF images and calculating protein abundance using standard curves for each staining batch.

### Annotation of regions of interest (ROIs) by the pathologist

After the image preparation, a board-certified pathologist (DLR) examined and circled the regions of interest (ROIs) of invasive tumor regions on pseudo-DAB images of individual cases in QuPath software (version 0.4.2). Regions of interest include only invasive cancer excluding normal tissue or stroma and selected to be qualitatively representative of the entire specimen. To assist in the ROI selection, we also performed H&E staining on each section adjacent to QIF-stained slides of individual cases for use when needed by the pathologist. The quantitative measurements of HER2 target are only in the ROIs defined by the pathologist.

### HER2 Quantitation in QuPath with QyMia extension

A molecular compartmentalization software extension for QuPath called “Qymia” was developed in our lab (Robbins et al)^18^. Qymia performs molecular compartmentalization analyses of regions of interest on whole tissue slides and tissue microarrays. The workflow allows users to label ROIs, define molecular compartments of interest, calculate biomarker expression scores per ROI annotation, convert Qymia scores to protein abundance measurements, and export measurements for further analyses. The cytokeratin immunofluorescence marker is used to define the tumor compartment within the pathologist annotated ROI based on intensity thresholding scripts performed in QuPath. Qymia’s approach is conceptually similar to AQUA software (McCabe et al)^19^ which uses molecular compartmentalization to quantify signal of biomarkers. For each annotated ROIs in selected BC cases, Qymia scores are proportional to the sum of the target pixel intensities within a molecular compartment divided by the area of the molecular compartment, 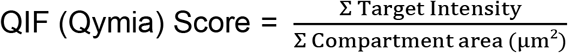.

### CMA Standard Curve Construction and Unit Conversion

In QuPath software with Qymia extension, the QIF images of control CMAs were used to measure HER2 signals in each cell lines by using TMA dearrayer and intensity thresholding QuPath scripts for the cytokeratin compartment. Then, Qymia scores were calculated for each CMA spot. CMA Qymia scores in each cell line were normalized by subtracting the average Qymia score of negative control cell line (Jurkat) spots on each CMA. The Qymia extension allows users to construct a standard curve by linear regression of normalized CMA Qymia scores by amol/mm^2^ abundance measurements of HER2 cell line spots. Supplemental figure 4 shows the standard curve of HER2 protein in the HER2 cell lines (Jurkat, BT-20, T47D, BT-483, ZR-75-1) from a single HS-HER2 run and a table of mass spec measurements of HER2 protein (amol/mm^2^) in each cell line. The linear regression equations of each CMA standard curve were used to convert the Qymia scores of HER2 protein amount in ROIs of core biopsies in BC cases which were stained in the same tray on Leica BOND RX into amol/mm^2^ units. The average of HER2 scores in cases with more than one ROI annotated by the pathologists were calculated in MS Excel.

### Determination of lower assay limits and statistical analysis

For the HS-HER2 assay, we verified the Limit of Detection (LOD) and Limit of Quantification (LOQ) according to guideline of the Q2(R1) Validation of analytical procedures: Text and Methodology-Guidance for Industry from the FDA. (https://www.fda.gov/regulatory-information/search-fda-guidance-documents/q2r1-validation-analytical-procedures-text-and-methodology-guidance-industry).^17^

The limit of detection (LOD) is defined as the lowest concentration of an analyte in a sample that can be consistently detected with a 95% confidence level while the limit of quantification (LOQ) is defined as the lowest amount of analyte in a sample which can be quantitatively determined with acceptable precision and accuracy. LOD & LOQ can be calculated using a series of calibration curves as follows.

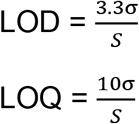

Where

- *σ* = standard deviation of y intercepts of the series of calibration curves
- S = average slope of the series of calibration curves

By using 6 CMAs from 6 independent experiments in HS-HER2 assay validation study, standard curve of average amol/mm^2^ values in HER2 low cell lines was used and we determined the limit of detection (LOD=360 amol/mm^2^) and limit of quantification (LOQ=1093 amol/mm^2^) to quantitatively analyze HER2 measurements in selected BC cases.

After determining lower assay limits of HS-HER2 assay, further statistical analysis was performed in GraphPad Prism v.10.2.0 (GraphPad Software Inc., CA, USA). Linearity of calibration curves was assessed using coefficient of determination (r^2^). Analysis of variance (ANOVA) for the correlation of baseline parameters with targets’ expression was performed, using F-test. Multivariable linear regression models were used to assess the impact of multiple parameters on targets expression. All statistical tests were two-sided with level of significance set at α<0.05.

## RESULTS

The first step for a quantitative analytic assay was to build a calibration standard^12^.Here our calibration standard used cell lines with target protein concentrations determined by mass spectrometry and converted to amol/mm^2^ as described previously (supplemental Figure 4). One standard slide was included with every 9 patient case slides in each tray of the autostainer. Since we measure on a continuous scale, we could not choose 20 “HER2 positives” and 20 “HER2 negatives”, as prescribed by the guidelines, but rather chose cases across the spectrum of HER2 expression (see Figure 1). To complete validation, we needed to compare this test to the legacy assay (clinically validated IHC assay at Yale). This is challenging since the legacy assay is less sensitive and ordinal compared to our more sensitive, continuous assay. We chose to compare this HS-HER2 assay to HER2 IHC by defining any case above the LOQ as positive and those below as negative to make the assay binary.

**Figure 1.**
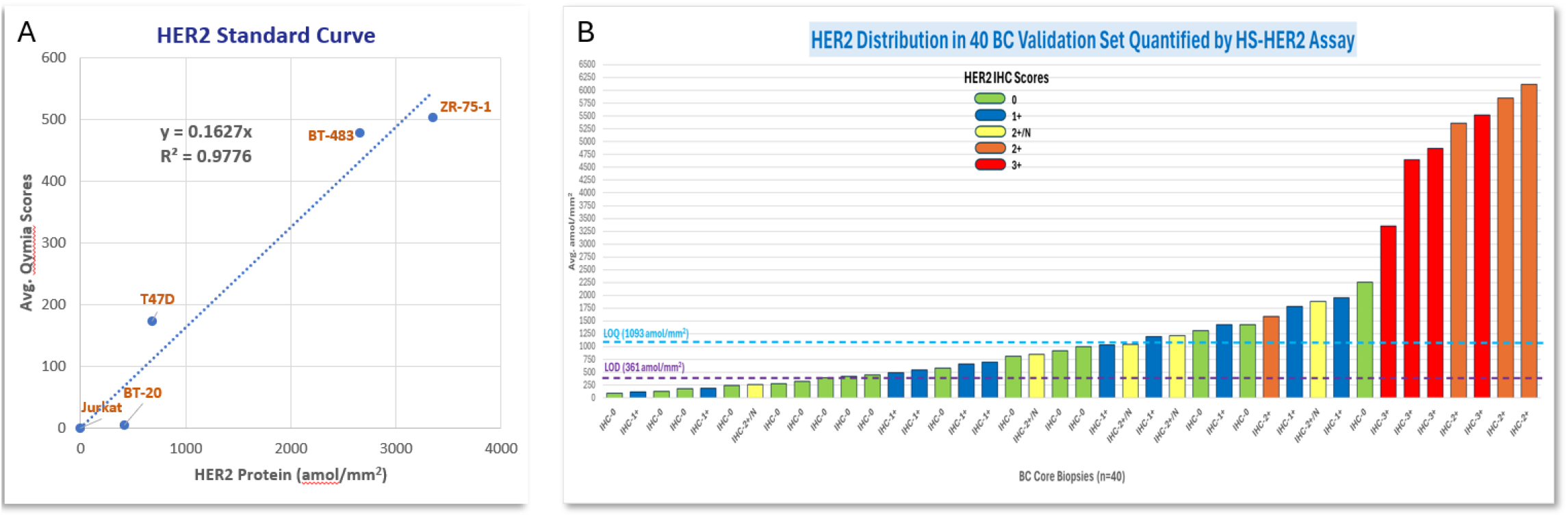
A) The standard curve used to convert fluorescent signal to amol/mm^2^ and B) the distribution of expression of the 40 validation set cases

We determined the LOD and LOQ using the mean values for the 6 standard curves used for the measurement of the validation study. Using this approach, for the validation set, our LOD was 361amol/mm^2^ and our LOQ was 1093 amol/mm^2^. The decision to use the LOQ is arbitrary in that there is no direct conversion between the continuous assay and the ordinal scoring system of the legacy assay and the cut-point, in amol/mm^2^, associated with response to either Trastuzumab or Trastuzumab Deruxtecan is unknown. Compared to the legacy HER2 IHC assay on the validation set cases the HS-HER2 sensitivity was 100%. However, since the HS-HER2 assay is more sensitive than the legacy assay some of the cases that were above the LOQ and defined as positive were negative by the legacy assay. Thus, specificity is 72% since it is calculated by categorizing our cases above, but near the LOQ as “false positive” since we call tests “positive” when they are negative by the legacy IHC assay.

Accuracy (sensitivity and specificity) is the main issue addressed in IHC validation guideline^13^as well as several technical issues. However, since the HS-HER2 assay is in some ways more similar to a truly objective analytic assay like the LBA (Ligand Binding Assay), we have also expanded our validation to include precision and reportable range. Analytic assay precision is assessed as the coefficient of variation (%CV) calculated as the standard deviation divided by the mean times 100. For HS-HER2 we calculated inter-assay precision using a protocol roughly adopted from the LBA approach. Three slides were run on 3 separate days with a minimum 1 week wash out period in between. The inter-assay %CV for our 40-case validation set was 10%. We also calculated intra-assay precision and inter-operator precision which were 6.4% and 7.3% respectively.

Reportable range is another variable that can determined for analytic assays but not IHC. In the M10 Bioanalytic Method Validation Guidance for Industry^20^, the reportable range for an analytic assay is defined by the lower LOQ (LLOQ) to the upper LOQ (ULOQ). Here we find the LLOQ for HS-HER2 to be 1093 attomol/mm^2^. While the ULOQ is more difficult to determine and for some assays is defined by the limit of linearity. Based on some preliminary response data, we have found direct proportionality between the amount of HER2 protein and the response to T-DXd, even above our calculated limit of linearity, and thus hesitate to define the limit of linearity as the upper end of the reportable range. Thus, we tentatively define the upper limit of our reportable range to be the highest expressing case in our 316 cases prospective trial (Figure 2) at 9434 amol/mm^2^.

**Figure 2.**
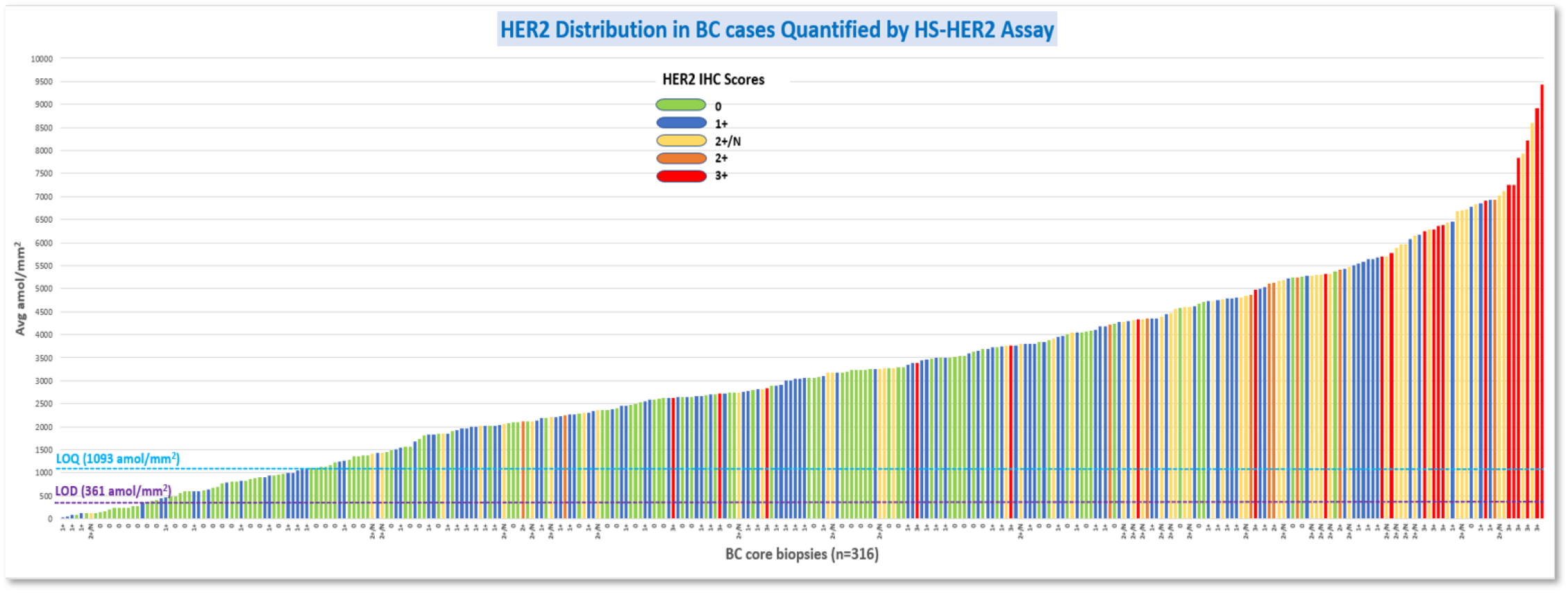
A histogram of 316 measurements of HER2 from the breast cancer core biopsy cases in the prospective trial sorted from lowest to highest and color coded by IHC scores (2+/N means a score of 2+ with reflex FISH testing showing no *ERBB2* gene amplification). The LOD and LOQ values for this assay are inset in the lower left.

In our pilot study of this HS-HER2 assay, our comparison of amol/mm^2^ showed very poor correlation with the IHC scores from the same cases (Moutafi *et al*.*12*). However, that work was limited by the fact that the IHC scores were done on whole tissue sections while the quantitative analysis was done on TMAs. To address that issue and to further validate the HS-HER2 assay, we designed and completed a prospective assessment of 316 breast cancer core biopsies received in the Yale Pathology Labs surgical pathology suite between August 2022 and August 2023. A total of 322 cases were collected of which 316 could be measured. Figure 2 shows the distribution of these cases sorted by amol/mm^2^ from lowest to highest color coded by the HER2 IHC scores retrieved from the Yale Pathology laboratory information system for each case. Again, there was a good concordance between concentration and high IHC scores, but IHC scores of 2 or below appear to be almost randomly distributed. Table 1 summarizes the level of expression by HS-HER2 compared to IHC scores. Notably, 71% of the IHC=0 cases and 84% of the IHC=1 cases were above the LOQ. Only 6% of the cases were below the LOD. Figure 3A and 3B illustrate the broad levels of measured HER2 expression in each IHC score category. Assessment of measurements by hormone receptor status suggests that each category shows a broad dynamic range of HER2 expression with no statistical significance with 95% confidence interval between subgroups (Figure 3C).

**Table 1.** Comparison of HER2 measurements to IHC scores.

| HS-HER2 Assay | IHC 0 |  | IHC 1+ |  | IHC 2+/N |  | IHC 2+ & 3+ |  | amol/mm2 |  |
| --- | --- | --- | --- | --- | --- | --- | --- | --- | --- | --- |
| Below LOD | 12 | 11% | 5 | 4% | 1 | 2% | 0 | 0% | 18 | 6% |
| Between LOD & LOQ | 21 | 19% | 14 | 12% | 0 | 0% | 0 | 0% | 35 | 11% |
| Above LOQ | 79 | 71% | 99 | 84% | 54 | 98% | 31 | 100% | 263 | 83% |
| Total Slides | 112 |  | 118 |  | 55 |  | 31 |  | 316 |  |
| Total Cases | 316 |  |  |  |  |  |  |  |  |  |

**Figure 3.**
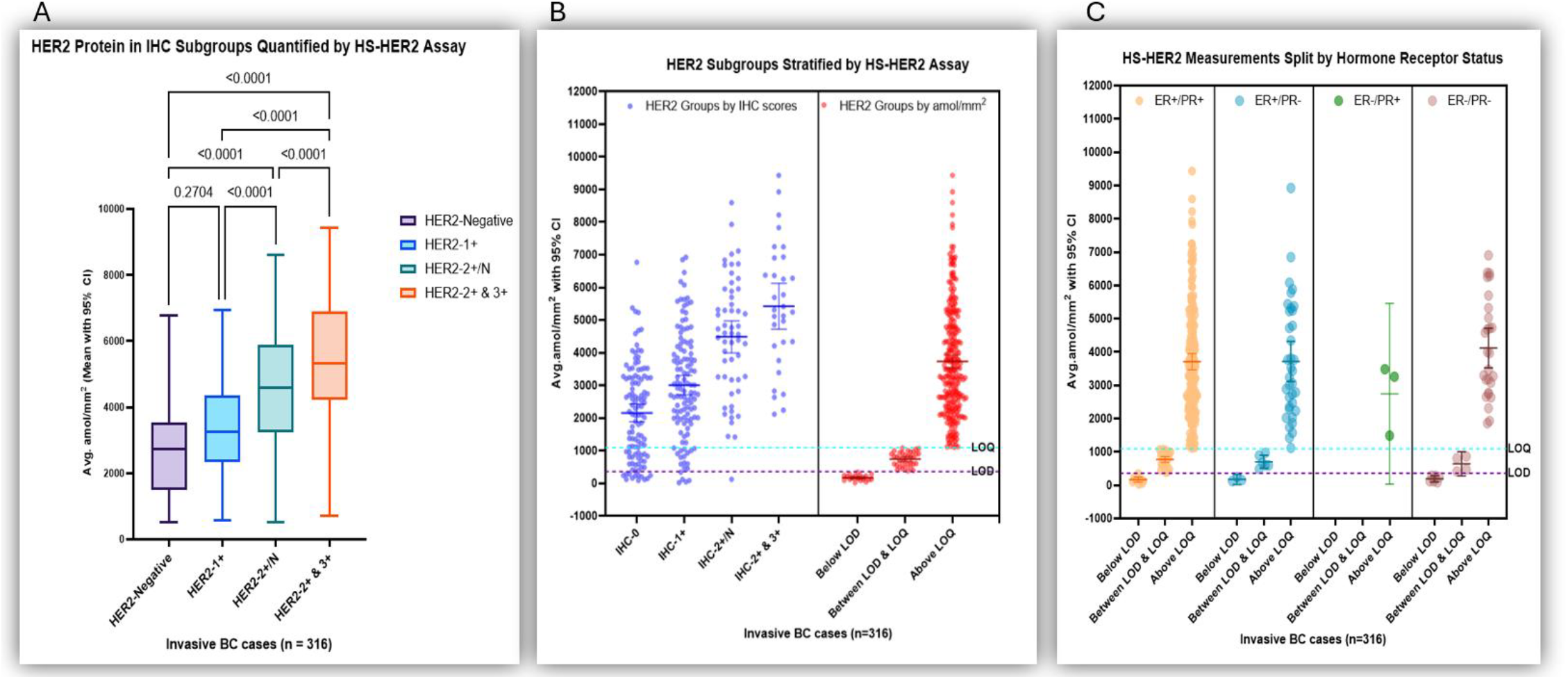
A) Box plot showing the HER2 proteins in IHC subgroups quantified by HS-HER2 assay; B) HER2 measurement (amol/mm^2^) in different subgroups stratified by traditional IHC scores (left) and by HS-HER2 assay limits (right); C) Distribution of HS-HER2 measurements split by hormone receptor status

## DISCUSSION

Assessment of the expression level of HER2 has become an important issue for optimizing patient care in the era of HER2 targeting ADCs. The DESTINY Breast-04 and -06 trials have changed to requirements for pathologists’ assessment, first requiring splitting of the ASCO/CAP “negative” category (IHC=1+ or IHC=0) then further splitting the IHC=0 category into IHC=0 and IHC>0 but <1. While there is ample published evidence that these new splits cannot be reproducibly read by pathologist^9-11,17,21-32^ even with training^33^, somehow the FDA approved both assays using an IHC test known to be in the wrong dynamic range for assessment of low level HER2 expression^34^. Careful examination of the Summary of Safety and Effectiveness Data documentation (SSED) from the FDA^35^shows that the FDA or the submission to the FDA used a disingenuous/dishonest method to reach the concordance levels needed for FDA standards for reproducibility.

Perhaps the best work in the literature for assessment of pathologists’ ability to reproducibly read HER2-Low comes from Ruschoff et al for the HER2-low Study Group in Archives of Pathology^34^The Study Group includes 77 pathologists from 14 countries. Table 1 in that paper shows that the overall rater agreement (ORA) for the 4B5 PATHWAY assay that is FDA approved is 83.9% for IHC=0 and 98.9% for IHC=3. Neither is significantly improved by training. The SSED assesses precision (reproducibility) by combining the IHC=3 with the IHC=0 in one category and the IHC=1 and =2 in the other category. This is not how it works in the real world. However, using this scheme, combining the task at which the pathologists are the best with the task at which we are not so good dilutes the discordance sufficiently that the SSED shows 95% or greater overall percent agreement (for both HER2-low and -ultra low). We interpret this disingenuous method as a cry for a better assay.

Here we describe fully quantitative and calibrated method for measurement, rather than reading of HER2. Our goal was to remove as much subjectivity as possible and make the assay more similar to blood glucose than an IHC scoring system. As such we used technology and computation efforts that allow standardization and evaluation in a manner similar to those done in laboratory medicine, specifically an LBA. We find that our method results in highly accurate assessment of HER2 protein expression with precision determined by calculation of the coefficient of variation to be under 10%. Furthermore, consistent with previously described assessment of this assay on TMAs (Moutafi *et al*.^12^), we find that, in a prospective study in a real world academic medical center, a high percentage of cases called IHC=0 were above the LOQ and a few cases called IHC=1 or IHC=2 were below the LOD.

This effort has some limitations that need to be considered. Most significantly, we used cell lines as a calibration tool. This is a challenge, and before we had developed a standardized SOP with strict criteria for growth, harvesting, storage and shipping, our calibration standard showed great variation. A careful reader will notice that calculation of amol/mm^2^ in our initial studies were significantly lower than the current calibration CMAs used in this work. We believe that with meticulous care, cell lines can be used as calibration standard, and we have included our CMA SOP as supplemental material to facilitate reproduction of this method by other groups. A second weakness is that this work represents a single institutional study. While our breast cancer pathologists carefully adhere to ASCO/CAP guidelines and the IHC reading for the 1-year prospective trial was done by 5 different pathologists, we realize that the work would be stronger if the cases were collected from multiple institutions and read by more than 5 pathologists. Finally, a limitation of this work is that it is hard to imagine that many labs would have the finances and equipment to replicate this work or offer this assay in their CLIA labs. We agree that it is complex but propose that the approach for clinical adoption be similar to Next Generation Sequencing (NGS). When NGS came along, it was clear that only the labs with high budgets, technical expertise and large catchment would be able to offer this service. We speculate that the HS-HER2 assay might see adoption in a pattern that is similar to NGS. Furthermore, we have begun efforts to transfer our academic CLIA lab protocols and technology to a large vendor in the anatomic pathology space.

## CONCLUSION

Herein we described a new quantitative, standardized assay to measure HER2. We have attempted to validate the assay in a hybrid manner, using requirements from both IHC and LBA protocols. Confirming previous work, in our prospective trial we found concerning levels of discordance between pathologists’ scores and measurements in amol/mm^2^. In light of the many ADCs in trials and likely future broad usage of this targeted therapy^1,36-40^ we believe that accurate assessment of the ADC targets will be critical for selecting the right drug for the right patient.

## Data Availability

All data produced in the present study are available upon reasonable request to the authors

## ETHICS DECLARATION

All tissue samples were collected under the approval of Yale Human Investigation Committee protocol #9505008219. Written informed consent, or waiver of consent, was obtained from all patients with approval of the Yale Human Investigation Committee. This work was conducted in accordance with the Declaration of Helsinki adopted by the world medical association.

## Conflict of Interest Statement

Dr. Rimm is a Consultant/Advisor to Astra Zeneca, Cell Signaling Technology, Cepheid, Danaher, Halda Therapeutics, Incendia, NucleAI, Regeneron, and Sanofi. Amgen, Astra Zeneca, Cepheid, NavigateBP, NextCure, Leica, Regeneron and Konica/Minolta currently fund, or have previously funded, research in his lab.

## ACKNOWLEDGEMENT

This work was funded by Yale Pathology Labs, a grant from the Breast Cancer Research Foundation (BCRF23-138) to DLR, and grants from the NIH including P30CA016359(DLR) and F30CA287869 (CJR).

## Notes

### Competing Interest Statement

Dr. Rimm has served as a Consultant/Advisor to Astra Zeneca, Cell Signaling Technology, Cepheid, Danaher, Halda Therapeutics, Incendia, NextCure, Nucleai, PAIGE.AI, Regeneron, and Sanofi. Astra Zeneca, Cepheid, NavigateBP, NextCure, Danaher/Leica, Regeneron and Konica/Minolta currently fund, or have previously funded, research in his lab.
Patricia Gaule is now an employee of BMS
Dr. Liebler is an employee and shareholder of Inotiv, Inc.
Regan Fulton is an owner of Array Science, LLC, and is a paid advisor to Leica Biosystems, AbbVie, and Personalis, Inc

## REFERENCES

1 Shastry, M. et al. Rise of Antibody-Drug Conjugates: The Present and Future. American Society of Clinical Oncology Educational Book (2023).

2 Modi, S., Saura, C., Yamashita, T. & et al. Trastuzumab Deruxtecan in Previously Treated HER2-Positive Metastatic Breast Cancer. N Engl J Med 382, 610–621 (2019).

3 Modi, S., Saura, C., Yamashita, T. & et al. Trastuzumab Deruxtecan in Previously Treated HER2-Positive Metastatic Breast Cancer: Updated Survival Results from a Phase II Trial (DESTINY-Breast01). Annals of Oncology 35, 302–307 (2023).

4 Modi, W., Jacot, T., Yamashita, T. & et al. Trastuzumab Deruxtecan in Previously Treated HER2-Low Advanced Breast Cancer. N Engl J Med 387, 8–20 (2022).

5 Bardia, A. & et al. Trastuzumab Deruxtecan after Endocrine Therapy in Metastatic Breast Cancer. N Engl J Med 391, 2216–2226 (2024).

6 Mosele, F., Deluche, E., Lusque, A. & et al. Trastuzumab Deruxtecan in Metastatic Breast Cancer with Variable HER2 Expression: The Phase 2 DAISY Trial. Nat Med 29, 2110–2120 (2023).

7 Ximena, B.-N., Roberto, S. & et al. Selecting Patients with HER2-Low Breast Cancer: Getting Out of the Tangle. European Journal of Cancer 175, 187–192 (2022).

8 Fernandez, A. I., Liu, M., Bellizzi, A. & et al. Examination of Low ERBB2 Protein Expression in Breast Cancer Tissue. JAMA Oncol 8, 1–4 (2022).

9 Griggs, J. J., Hamilton, A. S., Schwartz, K. L. & et al. Discordance Between Original and Central Laboratories in ER and HER2 Results in a Diverse, Population-Based Sample. Breast Cancer Res Treat 161, 375e384 (2017).

10 Kaufman, P. A., Bloom, K. J., Burris, H. & et al. Assessing the Discordance Rate Between Local and Central HER2 Testing in Women with Locally Determined HER2-Negative Breast Cancer. Cancer 120, 2657e2664 (2014).

11 Farshid, G. et al. Development and Validation of a HER2-Low Focused Immunohistochemical Scoring System with High-Interobserver Concordance: The Australian HER2-Low Breast Cancer Concordance Study. Mod Pathol 37, 100535 (2024).

12 Moutafi, M., Robbins, C. J., Yaghoobi, V., Fernandez, A. I. & et al. Quantitative Measurement of HER2 Expression to Subclassify ERBB2 Unamplified Breast Cancer. Lab Invest 102, 1101–1108 (2022).

13 Fitzgibbons, P. L., Bradley, L. A., Fatheree, L. A. & et al. Principles of Analytic Validation of Immunohistochemical Assays: Guideline from the College of American Pathologists Pathology and Laboratory Quality Center. Arch Pathol Lab Med 138, 1432–1443 (2014).

14 Press, M. F. & et al. Assessment of ERBB2/HER2 Status in HER2-Equivocal Breast Cancers by FISH and 2013/2014 ASCO-CAP Guidelines. JAMA Oncol 5, 366–375 (2019).

15 Wolff, A. C. & et al. Human Epidermal Growth Factor Receptor 2 Testing in Breast Cancer: ASCO-CAP Guidelines Update. JCO 41, 3867–3872 (2023).

16 MacNeil, T. & et al. Antibody Validation for Protein Expression on Tissue Slides: A Protocol for Immunohistochemistry. Biotechniques 69, 460–468 (2020).

17 J., R. C., I., F. A., G., H. & et al. Multi-Institutional Assessment of Pathologist Scoring HER2 Immunohistochemistry. Modern Pathology 36, 100032 (2022).

18 Qymia in QuPath [computer software] (GitHub, 2023).

19 McCabe, A., Dolled-Filhart, M., Camp, R. L. & Rimm, D. L. Automated Quantitative Analysis (AQUA) of In Situ Protein Expression, Antibody Concentration, and Prognosis. J Natl Cancer Inst 97, 1808–1815 (2005).

20 M10 Bioanalytical Method Validation and Study Sample Analysis, Jan 2025 Update. (FDA, 2025).

21 Torlakovic, E. E. Fit-for-Purpose Immunohistochemical Biomarkers. Endocr Pathol 29, 199e205 (2018).

22 Cuadros, M. & Villegas, R. Systematic Review of HER2 Breast Cancer Testing. Appl Immunohistochem Mol Morphol 17, 1e7 (2009).

23 Thomson, T. A., Hayes, M. M., Spinelli, J. J. & et al. HER-2/neu in Breast Cancer: Interobserver Variability and Performance of Immunohistochemistry with 4 Antibodies Compared with Fluorescent In Situ Hybridization. Mod Pathol 14, 1079e1086 (2001).

24 Varga, Z., Noske, A., Ramach, C., Padberg, B. & Moch, H. Assessment of HER2 Status in Breast Cancer: Overall Positivity Rate and Accuracy by Fluorescence In Situ Hybridization and Immunohistochemistry in a Single Institution Over 12 Years: A Quality Control Study. BMC Cancer 13, 615 (2013).

25 Vincent-Salomon, A., MacGrogan, G., Couturier, J. & et al. Calibration of Immunohistochemistry for Assessment of HER2 in Breast Cancer: Results of the French Multicentre GEFPICS Study. Histopathology 42, 337e347 (2003).

26 Zhao, B., Wang, Y. & Xu, H. Concordance of Immunohistochemistry and Fluorescence In Situ Hybridization for Assessment of HER2 Status in Breast Cancer Patients in Xinjiang Autonomous Region, China. Int J Clin Exp Pathol 10, 10459e10466 (2017).

27 Pauzi, M. S. H., Masir, N., Yahaya, A. & et al. HER2 Testing by Immunohistochemistry in Breast Cancer: A Multicenter Proficiency Ring Study. Indian J Pathol Microbiol 64, 677e682 (2021).

28 Paradiso, A., Marubini, E., Verderio, P. & et al. Interobserver Reproducibility of Immunohistochemical HER-2/neu Evaluation in Human Breast Cancer: The Real-World Experience. Int J Biol Markers 19, 147e154 (2004).

29 Pfitzner, B. M., Lederer, B., Lindner, J. & et al. Clinical Relevance and Concordance of HER2 Status in Local and Central Testing - an Analysis of 1581 HER2-Positive Breast Carcinomas Over 12 Years. Mod Pathol 31, 607e615 (2018).

30 Bartlett, J. M. S., Going, J. J., Mallon, E. A. & et al. Evaluating HER2 Amplification and Overexpression in Breast Cancer. J Pathol 195, 422e428 (2001).

31 Castera, C. & Bernet, L. HER2 Immunohistochemistry Inter-Observer Reproducibility in 205 Cases of Invasive Breast Carcinoma Additionally Tested by ISH. Ann Diagn Pathol 45, 151451 (2020).

32 Umemura, S., Osamura, R. Y., Akiyama, F. & et al. What Causes Discrepancies in HER2 Testing for Breast Cancer? A Japanese Ring Study in Conjunction with the Global Standard. Am J Clin Pathol 130, 883e891 (2008).

33 Rüschoff, J. et al. Abstract HER2-13: HER2-13 Proficiency assessment of HER2-low breast cancer scoring with the Ventana PATHWAY 4B5 and Dako HercepTest HER2 assays and the impact of pathologist training. Cancer Research 83, HER2-13-HER12-13 (2023).

34 Rüschoff, J., Penner, A., Ellis, I. O., Hammond, M. E. H. & et al. Global Study on the Accuracy of Human Epidermal Growth Factor Receptor 2-Low Diagnosis in Breast Cancer. Arch Pathol Lab Med (2024).

35 Administration, U. Summary of Safety and Effectiveness Data (SSED) PMA P990081. (FDA, 2000).

36 Maecker, H., Jonnalagadda, V., Bhakta, S., Jammalamadaka, V. & Junutula, J. R. Exploration of the antibody-drug conjugate clinical landscape. MAbs 15, 2229101–2229101 (2023).

37 Liu, K. et al. A review of the clinical efficacy of FDA-approved antibody-drug conjugates in human cancers. Mol Cancer 23, 62–62 (2024).

38 Drago, J. Z., Modi, S. & Chandarlapaty, S. Unlocking the potential of antibody–drug conjugates for cancer therapy. Nature Reviews Clinical Oncology 18, 327–344 (2021).

39 Robbins, C. J., Bates, K. M. & Rimm, D. L. HER2 testing: evolution and update for a companion diagnostic assay. Nature Reviews Clinical Oncology (2025).

40 Yoon, J. & Oh, D.-Y. HER2-targeted therapies beyond breast cancer — an update. Nature Reviews Clinical Oncology 21, 675–700 (2024).

